# Assessment of health-related quality of life in type 2 diabetes mellitus at Moi County referral hospital, Taita Taveta county

**DOI:** 10.1101/2023.01.31.23285237

**Authors:** Dredah Wughanga Mwadulo, Mbindu Madhavi, Beatrice Nkoroi

## Abstract

Health-related quality of life is one’s perceived status of life in value systems and cultural settings where they dwell in and in relation to their aspirations, expectations, ideals and fears. Diabetes has always been seen as a disease of affluence but over the years it is increasingly becoming a problem in developing countries. Globally, approximately 1.5 million deaths that occurred in 2012 due to diabetes, 80% of the deaths occurred in the developing nations. There is an increasing problem of non-transmittable diseases in Sub-Saharan Africa region which has brought a change in lifestyles like smoking, physical inactivity and unhealthy diet. Such non-communicable diseases include cardiovascular diseases, neuropathies & renal disorders which are commonly associated as the complications for diabetes mellitus. Measuring the extent an illness has affected health is of significance to the care and management of those individuals with chronic diseases whose remedy is not likely and since they require long-term management and care. Quality of life has a multivariate approach as it highlights information on bodily health, emotional health, functional and social health spheres of an individual’s view of the extent their health has been affected. This measure, therefore, provides holistic care for an individual with diabetes mellitus. This study seeks to solely look at how the wellness of individuals with diabetes mellitus is affected. The research adopted a cross-sectional descriptive design. 165 respondents were selected through systematic random sampling. The researcher used logistic regression whereby patient’s characteristics like foot problem (aOR7.348; p=0.005) and numbness/pain in hands/legs/feet (aOR=0.155; p<0.001) were associated with low quality of life. Over half of the study participants 127 (77%) indicated that their health-related quality of life would be better without diabetes mellitus as depicted by a diabetes specific QoL mean score of-1.88 approximating to “much better” (−2). The overall mean ADDQoL AWI score (−4.48) indicated that the health-related quality of life of the patients was negatively impaired by type two diabetes mellitus. Among the 19 domain specific items, the study participants rated “sex life” as the most negatively impacted/ important (WI= -5.14). In conclusion, type two diabetes mellitus negatively affected the quality of life and diabetes neuropathy which featured as foot problem and numbness of hands/feet needs to be considered as a predisposing factor to low quality of life in diabetics. Care provided should go beyond the standard treatment that looks into how treatment, lifestyle & behavioral modifications affect daily functioning and well-being of patients thus focusing more on the individual patient’s context and ultimately improving the health-related quality of life of type two diabetes mellitus patients.

## Introduction

Diabetes mellitus is a long term, progressive, non-transmittable disease that ensues when the pancreas either cannot excrete sufficient insulin or when the human cells are insensitive to the insulin produced by the pancreas. Over the years, diabetes has been associated with lifestyle modifications like physical inactivity, unhealthy diet, and obesity which can lead to complications associated with cardiovascular risks and renal disorders. This, in turn, reduces the quality of life and life expectancy in people with diabetes (39).

Over 280 million people have diabetes mellitus globally with this statistic expected to increase to about 400 million by the year 2030, with the biggest population being in the developing countries. In every 5 people living with diabetes in developing countries, 4 of them are men and women who are breadwinners of their families (28).

The effects of Diabetes go beyond the individual as they also affect families, societies, national productivity and have socio-economic effects, which in the developing countries co-exist with other diseases like cardiovascular diseases (39).

The National Diabetes strategy 2015-2020 in Kenya indicates that the occurrence of diabetes in the adult population is 4.56% (750,000 persons) and 20,000 deaths annually. Moreover, this is likely not to be the true picture as 60% of individuals diagnosed with diabetes in Kenya go to health facilities with unconnected complaints (23).

The social, bodily and emotional facets of health of an individual that are affected by the person’s values, hopes, and experiences are highlighted by the measure of their quality of life. It is then of significance for health care workers to recognize the bodily, social and emotional domains impacted by a chronic illness such as diabetes (16). Furthermore, the negative impact it has on the social domain, life expectancy and academic performance, ultimately affects their quality of life which is decreased by micro-vascular plus macro-vascular complications (16).

Assessment of one’s well-being in the context of health is increasingly becoming a significant end result measure of treatment and health interventions in patients with long-term illnesses (11).

Current care of diabetic patients focuses on treatment and how drugs can prolong their life span leaving out on the impact diabetes has socially, emotionally and financially (16).

Health is completeness in the physical, emotional and social health and not merely the lack of an illness or disability. For an individual to be healthy in all these aspects their well-being needs to be evaluated by measuring improvement in the extent to which his/her wellbeing has been affected by an illness (39). Healthcare researchers and workers consider this assessment as an indicator of not only evaluating the effect of chronic diseases on the lives of patients but also in measuring the efficiency of health interventions and treatment programs (14). Furthermore, individuals with diabetes mellitus are susceptible to premature aging, physical inactivity and weight gain in addition to debilitating complications of diabetes mellitus, negatively affecting their quality of life (14).

A lot of the literature on this subject of interest is from advanced countries Vis a Vis in developing countries where little is known. In addition, there is better healthcare provision in advanced countries rather than in developing countries where disability due to diabetes is associated with other morbidities (34).

## Materials and Methods

### Study design

A cross-sectional descriptive research design was used to describe what is happening at the current moment or specific point in time. This design was adopted as it sought to highlight differences between or association among the variety of study subjects (diabetic patients). The independent variables included socio-demographic and health-related characteristics (patient characteristics) which the researcher sought to find out how they were associated with either low/ high health related quality of life (dependent variable) in type 2 diabetes mellitus patients.

### Study population

They included all the patients with T2DM on follow up at the outpatient diabetic clinic at Moi County referral hospital in Taita-Taveta County. The facility receives a minimum of 90 patients monthly in the out-patient diabetic clinic.

### Sample & sampling technique

Calculation of desired sample size was by use of the Yamane formula. The study adopted a confidence interval of 95% and a sampling error of 0.05. The population size (P) was approximately 281 quarterly a year and *e* is 0.05

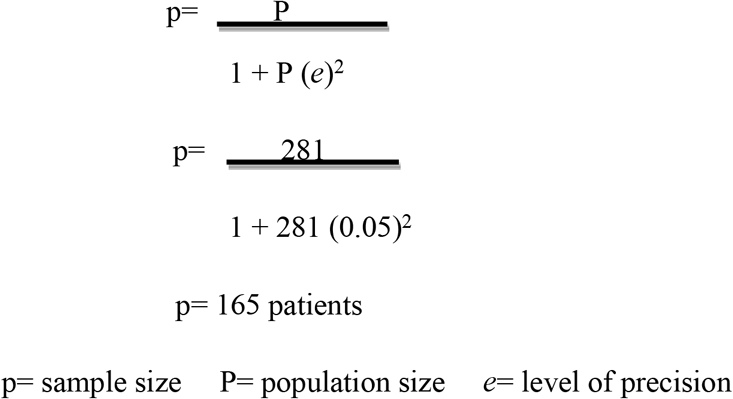

To cover for non-response, the sample size was be adjusted by using the following formula:

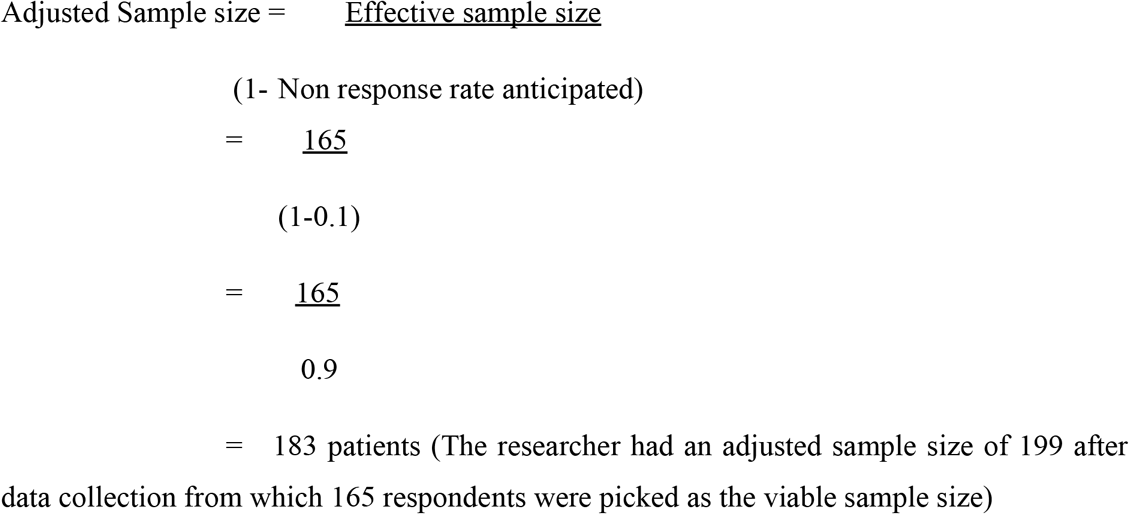

### Sampling technique

Systematic random sampling was used whereby every K^th^ patient was selected. In this research every 2^nd^ patient who came into the diabetic clinic was selected to be part of the sample size. The patients were assigned numbers and then randomly chosen by use of a sampling interval of 2. The sampling interval was considered by division of the population size by the desired sample size. The population size was **281** while the desired sample size was **165**. This is illustrated below:

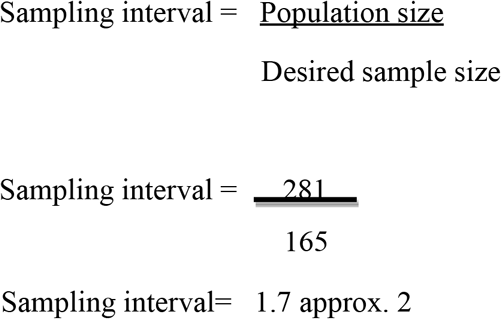

Therefore, every 2^nd^ patient was a member who was included in the sample size.

### Data collection tool

Quantitative data was collected by the use of a structured questionnaire and the ADDQoL-19 survey tool. The ADDQoL-19 which is a Patient-Reported Outcome Measure (PROM) was developed by Professor Claire Bradley for long-term health conditions like diabetes, chronic kidney disease, HIV and others (9). Professor Claire Bradley works at the Health Psychology Research limited in the United Kingdom (UK). The ADDQoL allows the respondent to rate only personally-applicable life domains, indicating importance and influence of diabetes.

The first two items of the tool assessed “present quality of life” and “diabetes dependent quality of life.” The first item range of scoring for “present quality of life” is from +3 (excellent) to -3 (extremely bad). The second item range of scoring for “diabetes dependent quality of life” is from +1 (worse) to -3 (very much better). This second item was evaluated by a scale from +1 (maximum positive impact of the diabetes) to -3 (maximum negative impact of the diabetes). The other 19-specific aspects of life followed. For example, item 1 and 2 of the tools covered leisure and work respectively. The impact of each specific aspect was evaluated by a scale from -3 (maximum negative impact of the diabetes) to +1 (maximum positive impact of the diabetes). The respondent then further stated how important this aspect is to their quality of life from a scale of +3 (very important) to 0 (not at all important) (9). Out of the 19-specific aspects of life, five had an optional response of “not applicable.” The weighted impact score for each aspect was calculated by multiplying the impact rating by the importance rating. The weighted impact score for each aspect was calculated by multiplying the impact rating by the importance rating. The average weighted impact score was calculated by adding the weighted evaluation of each aspect and diving it by the number of applicable specific aspects of life. The range of scoring the AWI was -9 (maximum negative impact of the diabetes) to +3 (maximum positive impact of the diabetes).

The structured questionnaire collected data on the patients’ characteristics (socio-demographic and health-related) while the ADDQoL survey tool measured the quality of life related to health of the diabetics on the aspects of bodily, emotional and social health domains. The structured questionnaire consisted of close-ended questions. Pre-test of the tool was done at Kilifi County referral hospital with 16 respondents which is 10% of the required sample size. Any unapplicable/ unclear questions in the structured questionnaire were either omitted or replaced.

### Data management and analysis

The quantitative data was processed by use of SPSS version 25. Descriptive statistics was used for the sociodemographic and health-related characteristics. Categorical variables were presented as percentages and frequencies while continuous variables were presented as mean and median. The researcher used bivariable logistic regression to assess the socio-demographic and health-related characteristics associated with the quality of life. All characteristics associated with quality of life in T2DM with a cut-off of P<0.25 in the bivariable analysis were considered for inclusion in the multi-variable logistic analysis. Furthermore, patient characteristics (socio-demographic & health related) in the multivariable logistic regression that had a P<0.05 were considered to be statistically significant and thus associated with the quality of life.

### Ethical consideration

Ethical clearance to do the study was sought from the research committee at Moi County Referral hospital through a letter from the Ethics and Research committee of Mount Kenya University. Also, clearance was sought from NACOSTI. The research license number from NACOSTI is NACOSTI/P/22/18413. A written consent was obtained from the study participants.

The researcher sought approval from the study participants prior to data collection through a written informed consent. The Informed consent was signed by all study participants. Anonymity was guaranteed for all participants. No financial incentives were offered for participation in the study. This study was approved by the Ethics Committee at Moi County Referral hospital. This study was approved by NACOSTI (research license number NACOSTI/P/22/18413). The ADDQoL-19 survey tool was adopted from Professor Claire Bradley of the Health Psychology Research limited in the UK. Permission to adopt the tool was sought and a license to use the tool in the research was issued which was reference number HPR 4458.

## Results

A total of 199 participants filled the research tool out of which 165 participants were identified as the required sample size. Most of the participants were female at 116 (70%), married were 127 (77%) and 114 (69%) had no source of income. Those living with diabetes for a duration of between 6 months-10 years were 113 (69%), most common treatment regimen was oral medication at 145 (88%) and co-morbidities like high blood pressure 113 (69%), eye/ vision problems 76 (46%), numbness or pain in feet/ legs 118 (72%) were the most common among the participants. This is illustrated in tab 1.

**Table 1:**
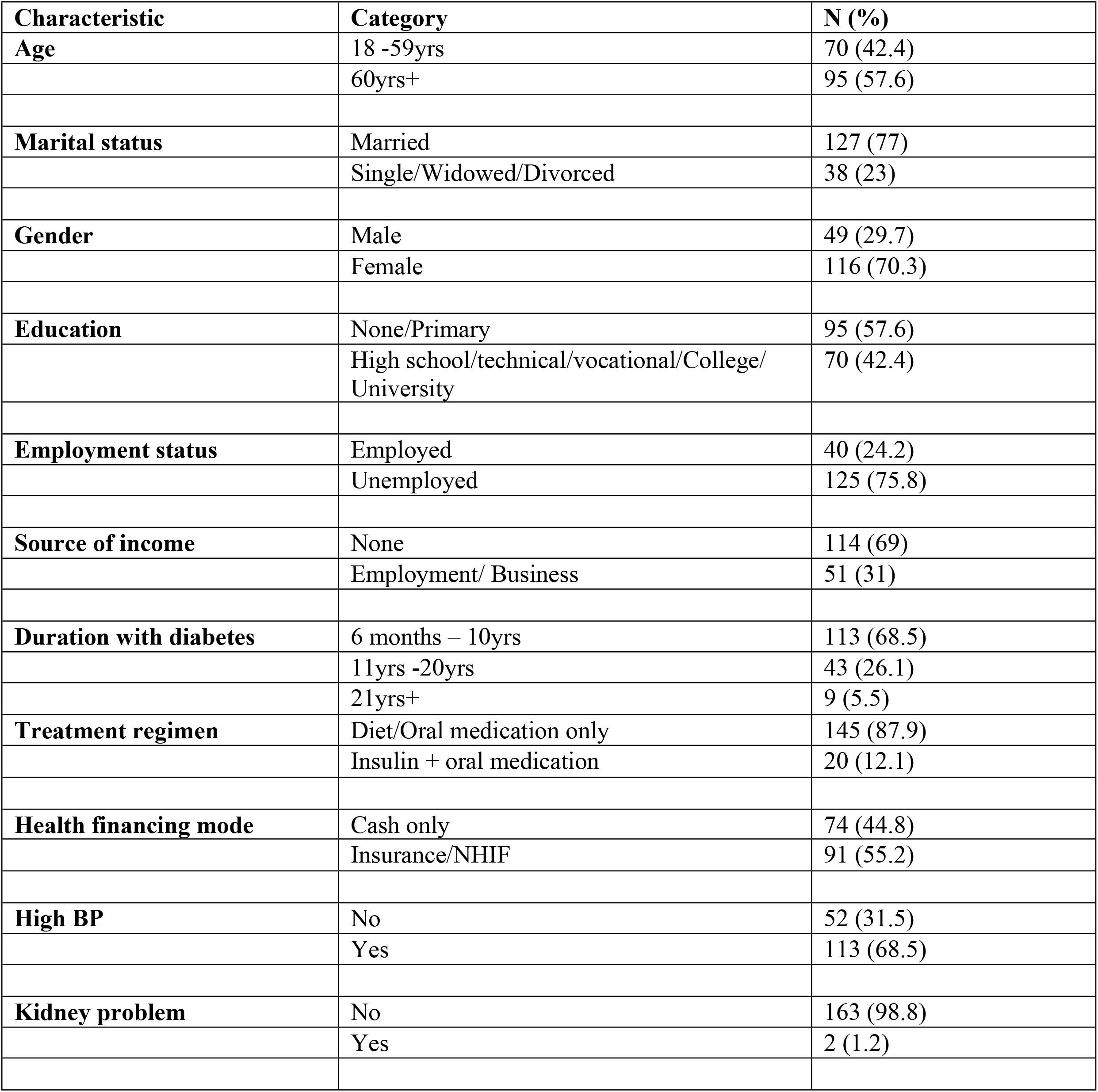
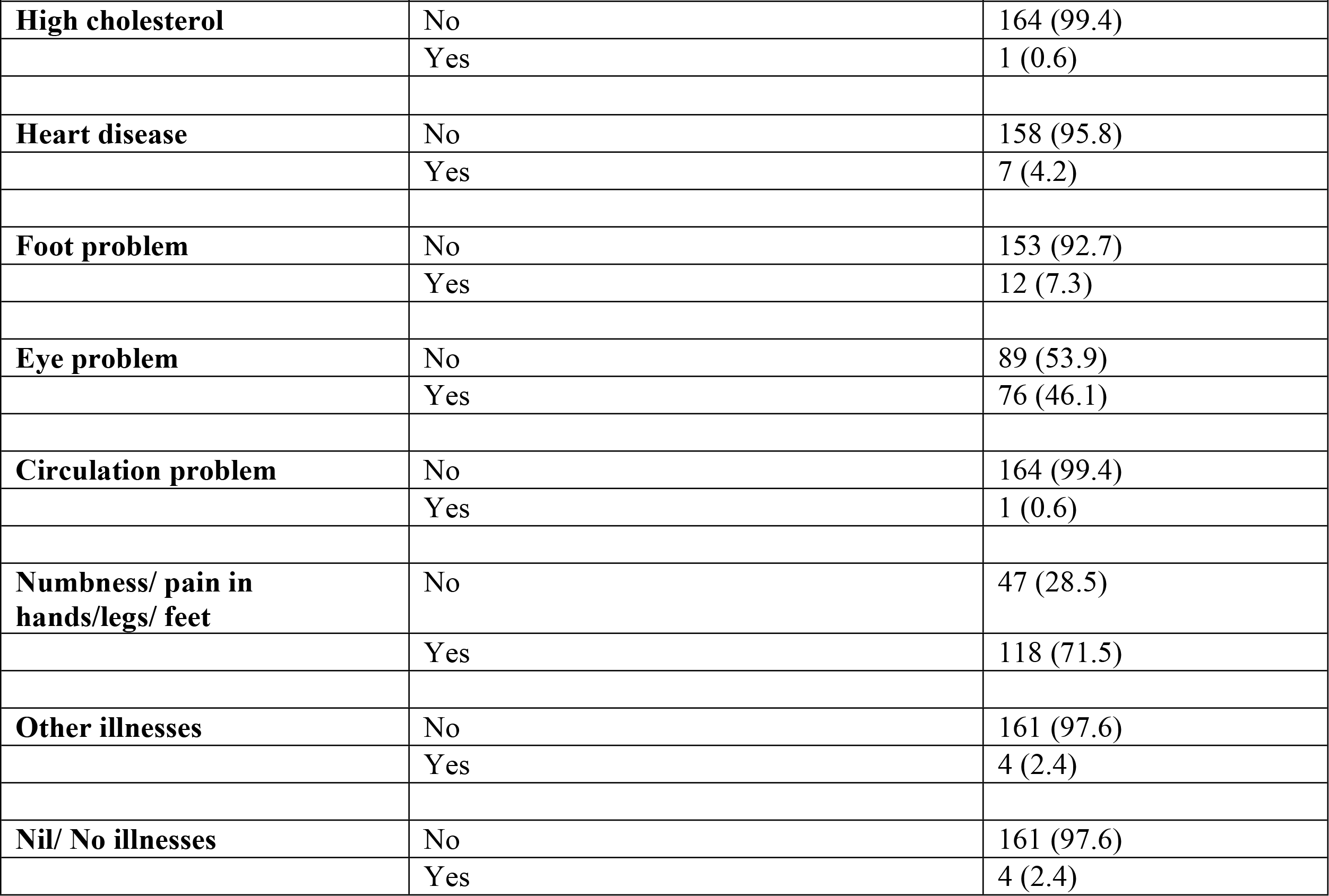
Distribution of study participants by socio-demographic and health-related characteristics (N=165)

In multivariable logistic regression a cut-off of P<0.25 was used where by the patient characteristics of duration of living with diabetes mellitus, co-morbidities like high blood pressure, foot problem and numbness/ pain in hands or feet or legs were analyzed. Patient characteristics that remained statistically significant after controlling other variables included foot problem and numbness/ pain in hands or legs or feet. Foot problem was at (aOR=7.348; 95% CI 1.824-29.605; P= 0.005) while numbness/ pain in hands or legs or feet was at (aOR= 0.155; 95% CI 0.062-0.389; P<0.001). This is illustrated in tab 2.

**Table 2:**
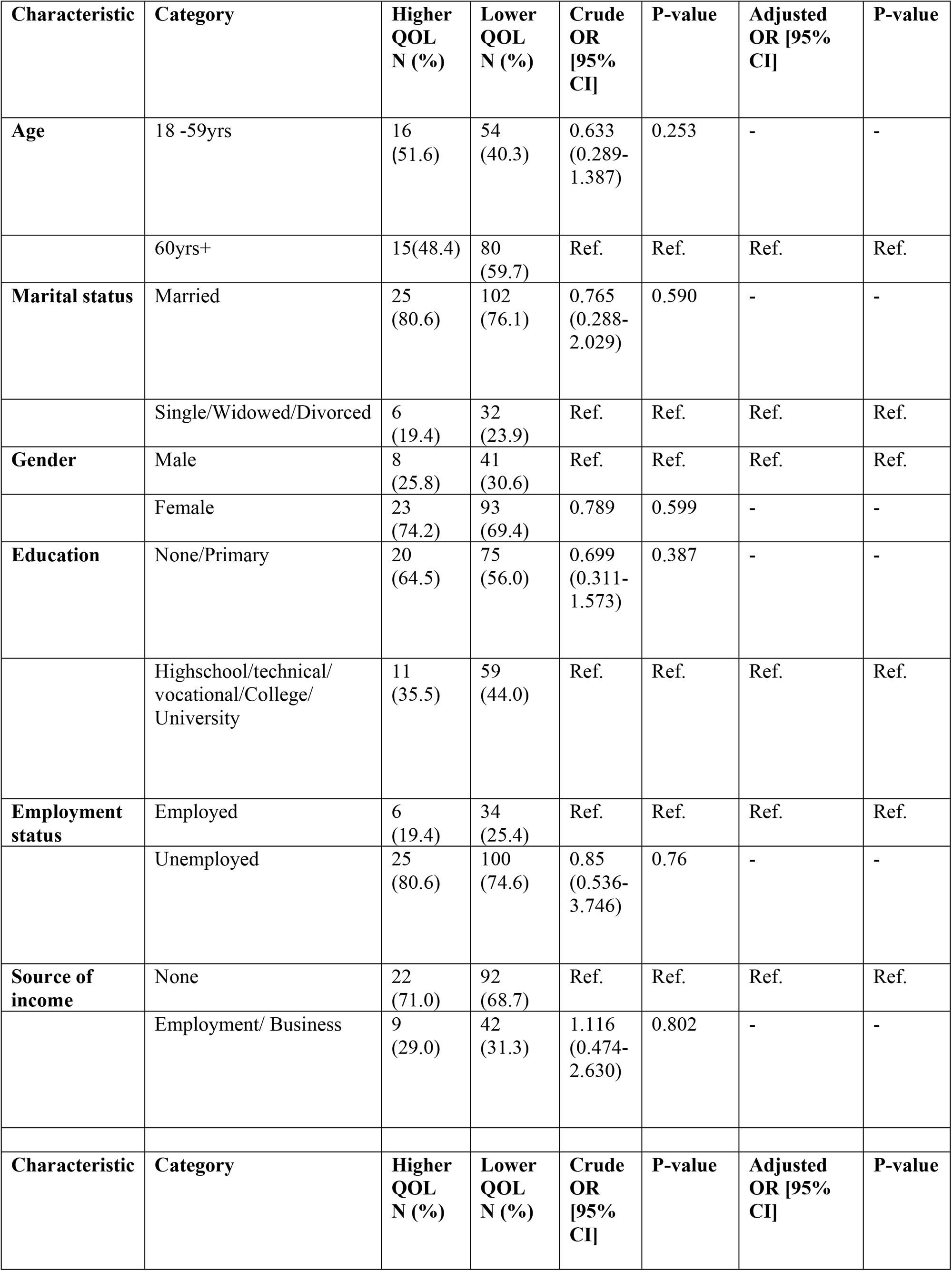
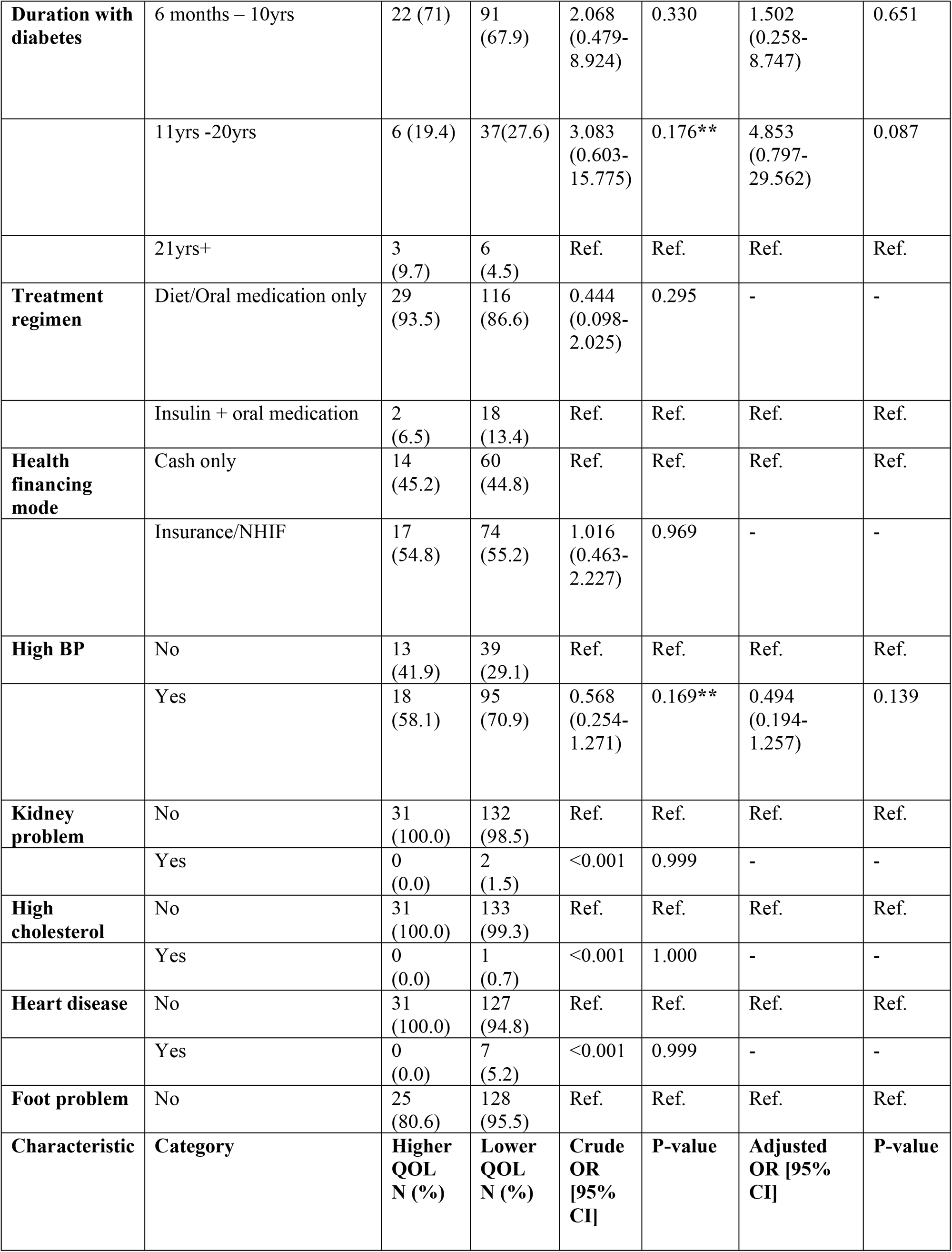
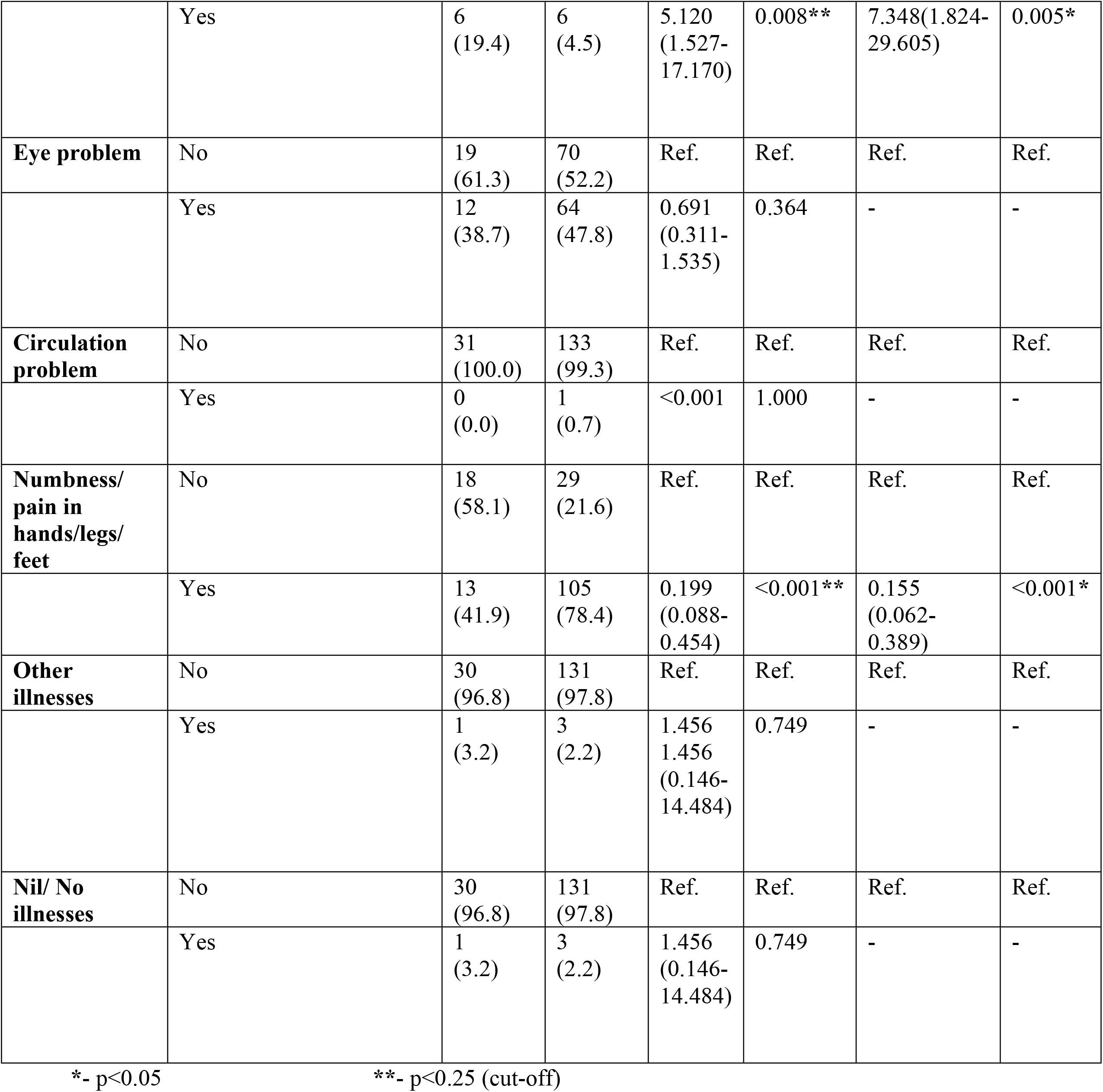
Socio-demographic and Health-related characteristics Multivariable analysis (N=165)

Over half 108 (65%) of the study participants stated that their quality of life was “neither good nor bad” while the least 3 (2%) stated that it was “bad” (tab 3).

**Table 3:** Responses to Overview Item one “perceived present quality of life” and overview item two “quality of life in the absence of diabetes mellitus”

| Particulars | Response | Frequency (N=165) | Percentage (%) |
| --- | --- | --- | --- |
| **Overview Item One: Perceived Present quality of life** | Bad | 3 | 1.8 |
|  | Neither good nor bad | 108 | 65.5 |
|  | Good | 48 | 29.1 |
|  | Very good | 6 | 3.6 |
|  | **Total** | **165** | **100.0** |
| Particulars | Response | Frequency (N=165) | Percentage (%) |
| **Overview Item Two: My quality of life in the absence of Diabetes mellitus** | Very much better | 10 | 6.1 |
|  | Much better | 127 | 77.0 |
|  | A little better | 26 | 15.8 |
|  | The same | 2 | 1.2 |
|  | **Total** | **165** | **100.0** |

Over half 127 (77%) of the study participants stated that their quality of life would be “much better” if they did not have diabetes mellitus while the least 2 (1%) stated that their quality of life would be “the same” if they did not have diabetes mellitus (tab 3).

The range of scoring of overview item one of the ADDQOL tool is -3.00, 3.00. The score per response was as follows: “excellent” (3), “very good” (2), “good” (1), “neither good nor bad” (0), “bad” (−1), “very bad” (−2), “extremely bad” (−3).

Overview item one of the ADDQOL-19 tool which looked into the present quality of life of the participants, the responses gave a mean generic quality of life score of 0.35 approximating to “neither good nor bad’’ (0) as opposed to “good” (1) or “very good” (2). This is illustrated in tab 4.

**Table 4:**
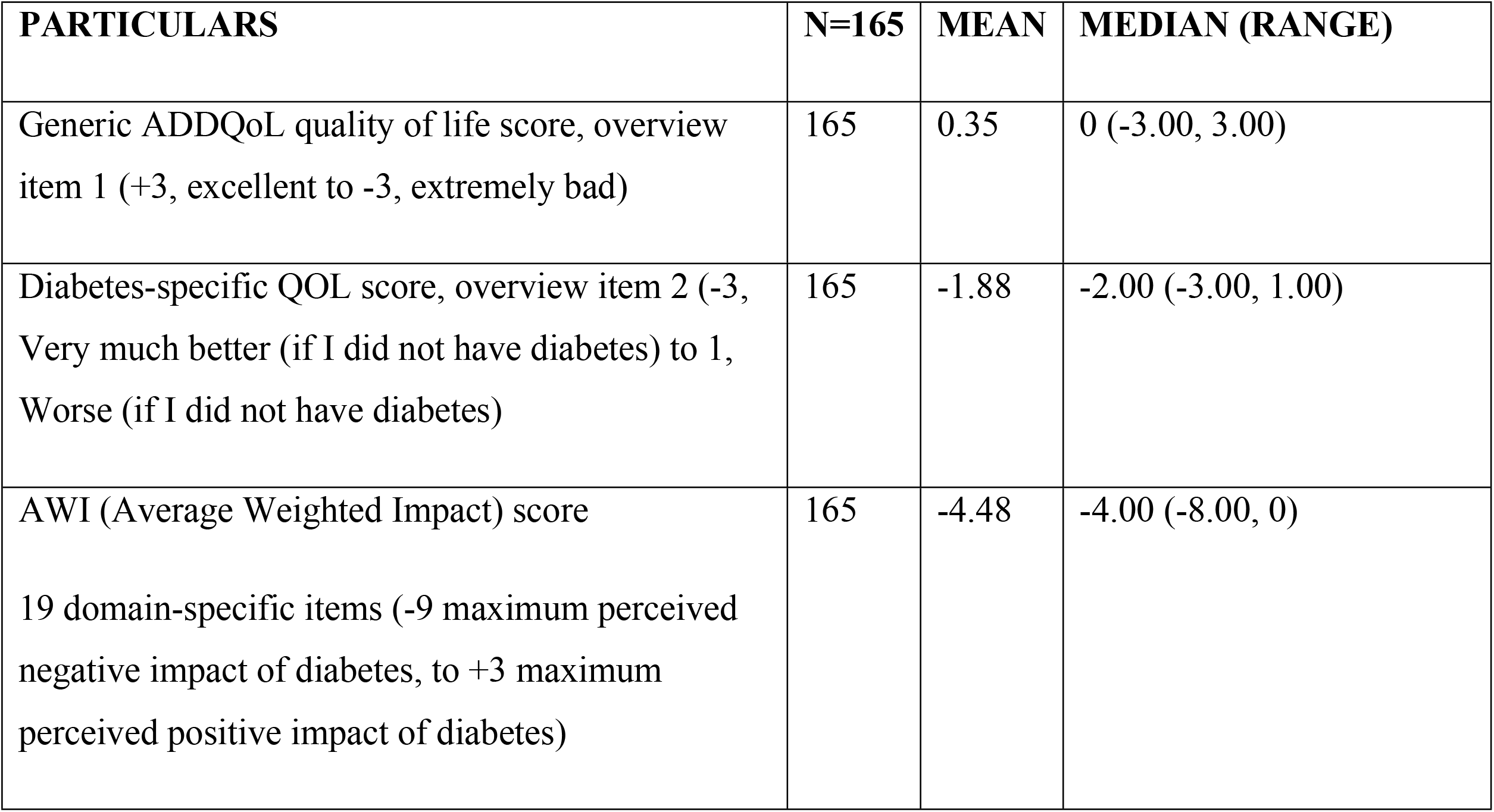
Mean and Median of Patient reported outcome measures for Generic quality of life and Perceived impact of diabetes mellitus type 2 on QoL

The range of scoring of overview item two of the ADDQOL tool is -3.00, 1.00. The score per response was follows: “very much better” (−3), “much better” (−2), “a little better” (−1), “the same” (0), “worse” (1).

Overview item two of the ADDQOL-19 tool looked into the perceived quality of life in the absence of diabetes mellitus. The responses gave a mean generic quality of life score of -1.88 approximating to “much better” (−2). This indicated that the health-quality of life would be better without/ in the absence of diabetes mellitus. This is illustrated in tab 4.

The mean AWI score for the domain specific items for the 165 study participants was -4.48, indicating a considerable perceived negative impact of diabetes mellitus type 2 on health-related quality of life. This is illustrated in tab 4.

The weighted impact score per domain-specific item was calculated by multiplying the impact rating by the importance rating. The range of scoring the weighted impact score was (−9.00) “maximum negative impact of diabetes mellitus” to (+3.00) “maximum positive impact of diabetes mellitus.” Among the 19 domain-specific items, the study participants rated “sex life” as the most negatively impacted (WI= -5.14) significantly more so than other items. This is illustrated in tab 5.

**Table 5:**
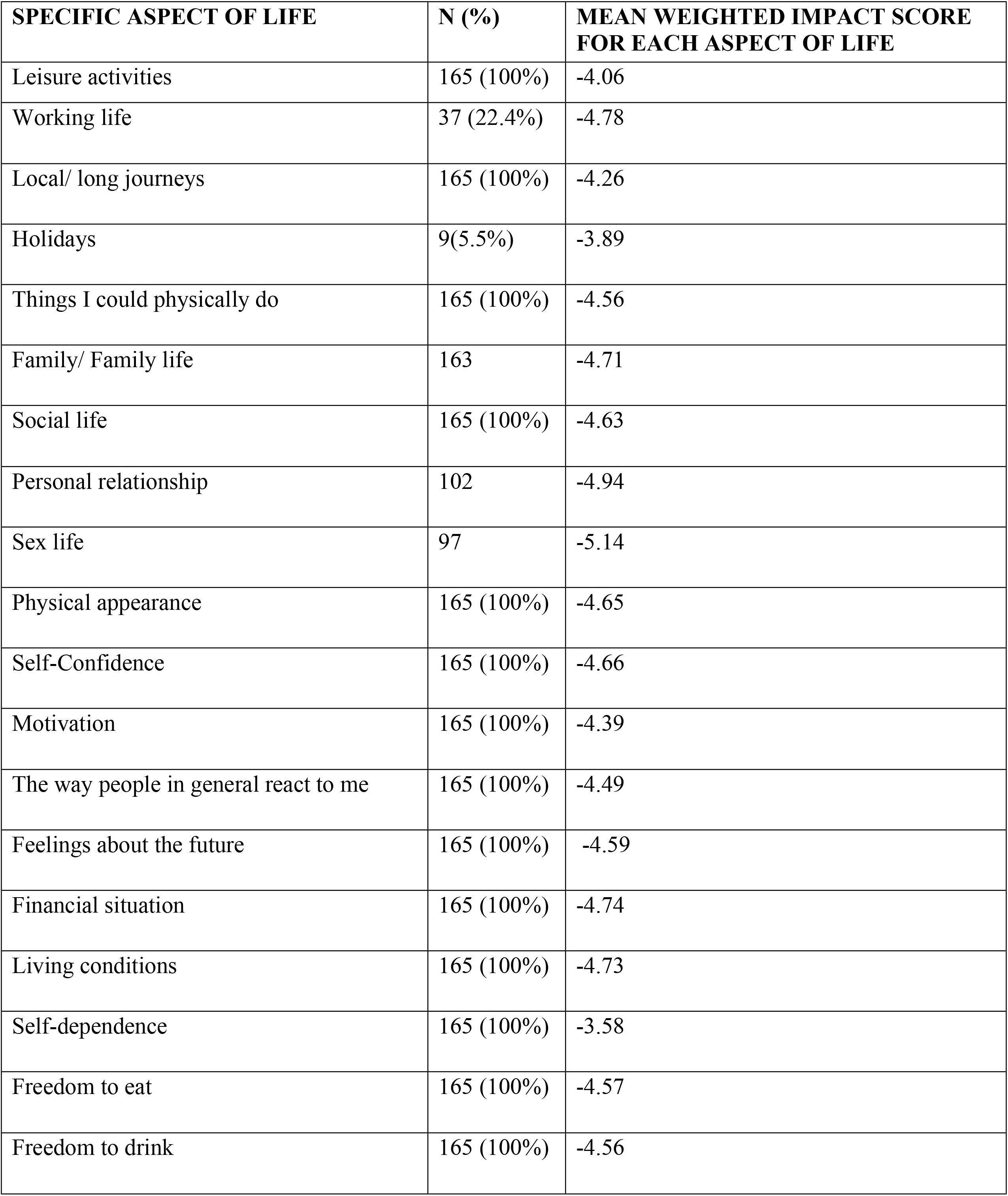
Mean ADDQoL Weighted impact item score

## Discussion

The patient characteristics that were statistically significant after multivariable logistic analysis was, foot problem and numbness/pain in hands/legs/feet. The research findings indicated that foot problem and numbness in hands/ legs/feet were the health-related characteristics related to low HRQOL among type two diabetes mellitus patients. Study participants who had foot problem were seven times more likely to have low health-related quality of life compared to those who did not have the comorbidity. In addition, study participants who had numbness/pain in hands/legs/feet were more likely to have low health-related quality of life compared to those who did not have the co-morbidity. These study findings are similar to a study done in Brazil by (22) that showed that 34 (17%) of the elderly diabetic participants who had foot wounds had lower quality of life due to decreased autonomy and social participation as a result of decreased mobility associated with neuropathic/vascular pain. Similar study findings were found in Saudi Arabia (3) which showed that despite the overall health related quality of life being moderate, 69% of the study participants who had pain/discomfort had severe to extreme health state that translated to low health-related quality of life (3). On the influence of type two diabetes mellitus on the health-related quality, the research findings indicated a considerable perceived negative impact of diabetes mellitus type 2 on the HRQOL of the 165 patients. Over half of the study participants perceived that their health-related quality of life would be better without/ in the absence of type two diabetes mellitus. This research finding is similar to a Bulgarian study which showed that 275 (67.1%) of the respondents perceived that their quality of life would be better without T2DM since the diabetes specific QoL mean score was at -1.8 (5).

The ADDQoL scoring ignores non-applicable domains and gives greater emphasis to domains of greater importance to the individual. Among the 19 domain specific items, the study participants rated “sex life” as the most negatively impacted/ important as compared to other items. This is despite also all other items being negatively influenced by T2DM. This was further coupled with negative influence of T2DM on the health-related quality of life which was indicated with a negative mean score of the AWI of the 165 study participants. This research finding is in contrast to most other studies using the ADDQoL, where the item reflecting the greatest negative influence of diabetes on the health-related quality of life was “Freedom to eat as I wish.” A study (10) indicated that “freedom to eat as I wish” was the most negatively impacted at (WI= -3.35). Also, a study done in Bulgaria showed that “freedom to eat as I wish” was the most negatively impacted at (WI= -4.0) followed by “Family life” WI= -3.9. In addition, the mean AWI score for the population was -2.9 which indicated an overall negative impact of T2DM on quality of life (5).

## Conclusions

Over half of the study participants 127 (77%) indicated that their health-related quality of life would be better in the absence of diabetes mellitus (5). The overall mean ADDQoL AWI score (4.48) indicated that the health-related quality of life was negatively impaired by type two diabetes mellitus. Since over half of the study participants perceived a low health-related quality of life then it can be concluded that most were aware of the influence of T2DM on their health status and ultimately on their health-related quality of life (3).

Co-morbidities of foot problem and numbness/pain in hands/ legs/feet were associated with low health-related quality of life. This may be due to the fact that diabetes neuropathy and neuropathic pain is one of the many complications associated with diabetes mellitus (22). Sexual satisfaction/ sexual function was an item which was the most negatively impacted and most important domain of the patient’s quality of life while “self-dependence” was the least negatively impacted domain. This may be due to the fact that microvascular complications like sexual dysfunction, infertility/ impotence is associated with diabetes mellitus (5)(10).

## Data Availability

All relevant data are within the manuscript and its Supporting Information files.

## Acknowledgements

The researcher would like to acknowledge her research methodology lecturer Dr. Nilufar for instilling in me the most appropriate research skills and knowledge. Secondly, I thank my thesis supervisors Professor Bindu Madhavi and Ms. Beatrice Nkoroi whose continuous support has been instrumental in the development of my thesis. Lastly to my husband who has been giving me the essential psychological support throughout my thesis.

## Informed Consent

### Introduction

You are requested to take part in this study **on assessment of the quality of life related to health in diabetes mellitus type 2 patients**.

I request you to go through this consent form and if you have any questions before agreeing to take part in this study, do not hesitate to ask the researcher. Before you decide to take part in the study, it is important that you know the following:

a. Your participation in this study is voluntary and refusal to participate will not be used against you.
b. Information acquired from this study will be kept confidential and the researcher will ensure anonymity of the study participants.
c. You have a right to withdraw from the study at any level. Your decision will be respected without consequences placed on you.

### What will happen if you decide to take part in this research study?

If you agree to participate in this study, you will be interviewed by the researcher in a private area/ area of your convenience where you will feel comfortable while answering the questions. The interview will last approximately 30 minutes.

The researcher will ensure confidentiality of the information given and anonymity of the study participant.

### Potential benefits and importance of the knowledge gained

There are no direct benefits to you as a study participant in this study.

The information acquired from this study will help the researcher to advice Moi County referral hospital on care of patients with diabetes mellitus type two.

If you have questions about this study, contact:

Principal Researcher

Dredah Mwadulo on mobile no 0726684128 or

## CONSENT FORM (STATEMENT OF CONSENT)

### Participant’s statement

I have read this consent form / The information on this consent form has been read to me.

I have had my questions answered in a language that I understand. I understand that my participation in this study is voluntary and that I may choose to withdraw any time. I agree to take part in this research study. I understand that all efforts will be made to keep information regarding my personal identity confidential.

Participant signature:_________ Date:__________

